# The relationship between relative aerobic load, energy cost, and speed of walking in individuals post-stroke

**DOI:** 10.1101/2021.03.22.21253569

**Authors:** Ilse Blokland, Arianne Gravesteijn, Mathijs Busse, Floor Groot, Coen van Bennekom, Jaap van Dieen, Jos de Koning, Han Houdijk

**Affiliations:** Department of Human Movement Sciences, Faculty of Behavioural and Movement Sciences, Vrije Universiteit Amsterdam, Amsterdam Movement Sciences, The Netherlands; Heliomare Research and Development, Wijk aan Zee, The Netherlands; Coronel Institute of Occupational Health, Amsterdam University Medical Center, University of Amsterdam, Amsterdam, The Netherlands; Sport- en BeweegKliniek, Haarlem, The Netherlands; University of Groningen, University Medical Center Groningen, Center for Human Movement Sciences, Groningen, The Netherlands

**Keywords:** Gait, Exercise Test, Oxygen consumption, Anaerobic Threshold, Cardiorespiratory Fitness (CRF), Physical Fitness

## Abstract

**Background:** Individuals post-stroke walk slower than their able-bodied peers, which limits participation. This might be attributed to neurological impairments, but could also be caused by a mismatch between aerobic capacity and aerobic load of walking.

**Research question:** What is the potential impact of aerobic capacity and aerobic load of walking on walking ability post-stroke?

**Methods:** In a cross-sectional study, forty individuals post-stroke (more impaired N=21; preferred walking speed (PWS)<0.8m/s, less impaired N=19), and 15 able-bodied individuals performed five, 5-minute treadmill walking trials at 70%, 85%, 100%, 115% and 130% of PWS. Energy expenditure (mlO_2_/kg/min) and energy cost (mlO_2_/kg/m) were derived from oxygen uptake 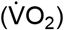. Relative load was defined as energy expenditure divided by peak aerobic capacity 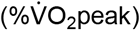 and by 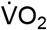 at ventilatory threshold 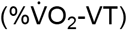. Relative load and energy cost at PWS were compared between groups with one-way ANOVA’s. The effect of speed on these parameters was modeled with GEE.

**Results:** Both more and less impaired individuals post-stroke showed lower PWS than able-bodied controls (0.44[0.19-0.76] and 1.04[0.81-1.43] vs 1.36[0.89-1.53] m/s) and higher relative load at PWS (50.2±14.4 and 51.7±16.8 vs 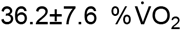 peak and 101.9±20.5 and 97.0±27.3 vs 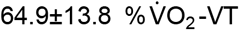). No differences in relative load were found between stroke groups. Energy cost at PWS of more impaired (0.30[.19-1.03] mlO_2_/kg/m) was higher than less-impaired (0.19[0.10-0.24] mlO_2_/kg/m) and able-bodied (0.15[0.13-0.18] mlO_2_/kg/m). For post-stroke individuals, increasing walking speed above PWS decreased energy cost, but resulted in a relative load above endurance threshold.

**Significance:** Individuals post-stroke seem to reduce walking speed to prevent unsustainably high relative aerobic loads at the expense of reduced economy. When aiming to improve walking ability in individuals post-stroke, it is important to consider training aerobic capacity.

## Introduction

Individuals post-stroke walk slower compared to able-bodied individuals, which can lead to problems in participation[1,2]. When addressing walking ability, underlying neurological factors such as reduced motor control, muscle weakness and spasticity, and concomitant biomechanical constraints, are often assessed. However, another possible reason for slow walking could be a high relative aerobic load caused by an increased energetic demand and a reduced aerobic capacity.

Aerobic capacity is often reduced in individuals post-stroke. On average, their peak capacity 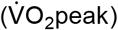 is about half that of able-bodied individuals[3]. In addition, their ventilatory threshold (VT), which represents the upper limit of workloads that can be sustained for a prolonged period[4], is below the 10^th^ percentile of able-bodied individuals[5–7]. Aside from a low aerobic capacity, individuals post-stroke experience a significantly higher aerobic load during walking (in mlO_2_/kg/min) than able-bodied peers[8]. This implies that the aerobic load relative to their aerobic capacity, i.e. the relative aerobic load of walking, is higher after stroke. Consequently, normal walking speed might require energy expenditure above sustainable limits. Hence, reducing walking speed might be a strategy to reduce relative aerobic load to sustainable levels.

Indeed, literature suggests that, at their lower preferred walking speed (PWS), individuals post-stroke experience aerobic loads similar to able-bodied peers[9–12]. This slow walking speed, however, can be problematic in daily life, for example when trying to keep up with a partner or when participating in traffic. This may cause avoidance of walking, causing a vicious cycle in which reduced physical activity leads to a further decline in fitness, aggravating mobility problems[13].

Aside from practical consequences, slow walking speed may have another disadvantage, namely an increased energy cost. The energy required per unit distance, generally referred to as energy cost, shows a U-shaped relationship with speed. The PWS of able-bodied people is generally close to the walking speed that requires the least amount of energy to cover a given distance[10,14]. Post-stroke however, people seem to walk below their most economic speed, aggravating the difference in energy cost compared to able-bodied[15,16].

Several authors have highlighted the importance of the limited aerobic capacity in relation to aerobic load during walking post-stroke[10,17,18], but only a limited number of studies reported relative aerobic load. Relative aerobic loads of walking in individuals post-stroke were between 45 and 65% of peak aerobic capacity 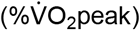 [19,20]. However, neither of these studies included a control group, nor did they impose speeds below and above PWS, to investigate the concomitant effect on relative load and energy cost.

The goal of this study was to identify to what extent physiological parameters could impact walking ability of individuals post-stroke. To this end, we compared relative aerobic load and energy cost at PWS between individuals post-stroke and able-bodied peers. Furthermore, we aimed to clarify the potential selection of PWS by investigating how these physiological parameters change as a function of walking speed. To assess the effect of stroke severity on these parameters, we stratified the stroke group into more and less impaired individuals based on PWS. We hypothesized that individuals post-stroke walk slower and experience a similar or higher relative aerobic load than able-bodied controls at PWS. We also hypothesized that in individuals post-stroke, PWS is below their most economic walking speed. Consequently, we expected that faster walking would reduce energy cost in individuals post-stroke, but at the expense of unsustainable relative load levels. We expected these effects to be larger in the more impaired individuals post-stroke.

## Methods

Forty-five people in the sub-acute phase after stroke[21] and fifteen age-, gender-, and BMI-matched able-bodied participants enrolled in this cross-sectional study. Individuals post-stroke were all in-patients in the rehabilitation center that were referred for a routine cardiopulmonary exercise test (CPET).

Inclusion criteria for individuals post-stroke were: (1) >18 years of age, (2) ability to sustain walking on a treadmill for a minimum of 4 minutes and (3) Functional Ambulation Category (FAC) >2, indicating ability to walk without manual assistance. Exclusion criteria for all participants were (1) contraindications for CPET and/or exercise[22,23]; and (2) inability to understand or perform instructions. Additionally, control participants were excluded if they had a history of stroke or heart disease, or if they had a disorder influencing walking ability. This study was approved by the Medical Ethical Committee of the VU Medical Center (NL-64431.029.18). All participants were fully informed about the study aim and protocol and signed written informed consent before participation.

### Procedures

Participants performed the CPET and treadmill walking protocol on two days scheduled 2 to 14 days apart. Participants were asked to refrain from heavy exercise 24 hours before the measurements and to refrain from taking food and caffeinated beverages for 2 hours before the measurements. Before testing, participants’ body height and mass were measured. Additionally for individuals post-stroke, the Motricity Index[24] and FAC[25] were determined. The following characteristics were retrieved from patient files: time since stroke, side and type of stroke, 1^st^ or recurrent stroke, days since admission, use of Beta Blockers and Berg Balance Score[26,27].

### Cardiopulmonary Exercise Test (CPET)

The CPET was performed on an electronically braked cycle-ergometer under supervision of an experienced physician. After a 3-min rest phase and a 3-min warming-up at 0 Watt, a ramp protocol based upon the estimated maximum exercise capacity of the participant was used. The ramp phase was stopped if the patient could not sustain a pace of 50 rounds per minute, if the physician deemed it unwise to continue the test, or if the participant wanted to stop. A cooling-down phase of 3 minutes at 10% of peak power followed.

Breath-by-breath gas exchange (Jaeger Oxycon Pro, Vyaire, Hoechberg, Germany) and arterial oxygen saturation were continuously measured. Additionally, for individuals post-stroke, a 12-lead electrocardiogram was monitored. Immediately after the ramp phase, rating of perceived exertion (RPE) was recorded on a modified Borg scale (6-20)[28].

### Treadmill Walking

The second measurement day started with a 10-minute, seated, resting oxygen uptake measurement. Subsequently, PWS was determined on the treadmill, using a protocol previously described[29]. Then, participants performed five treadmill-walking trials at different speeds in a randomized order. Each trial lasted five minutes and speed was fixed at either 70%, 85%, 100%, 115% or 130% of PWS. Seated rest between trials was at least five minutes to ensure return to resting metabolic values. Breath-by-breath gas exchange measurements and heart rate (Polar Wearlink strap, Kempele, Finland) were monitored continuously during the trials. RPE was recorded at the end of each walking trial.

### Data analysis

Peak aerobic capacity 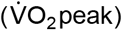 was determined from CPET data. The maximum of the 30-second averaged 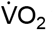 was considered 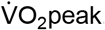. 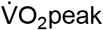 was considered valid when a participant reached two out of three following criteria: respiratory quotient >1.1 (>1.0 for participants over 65 years of age[30,31]), maximum heart rate >85% of predicted maximum, and RPE>15.

In addition to 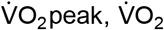 at ventilatory threshold 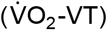 was used as an additional measure of aerobic capacity. 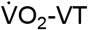 expresses the exercise intensity that can be sustained for long duration without involvement of anaerobic metabolism and has been suggested to be a more specific measure of endurance for individuals post-stroke[5]. Two assessors determined VT following the V-slope method[32]. A plot of the ventilatory equivalents and respiratory quotient over time was presented after V-slope assessment, to allow the assessor to adjust the time point of VT if needed. If the two assessors did not agree on the 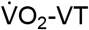 within 100 ml/min they conferred to reach consensus.

For the walking trials, the occurrence of a steady-state 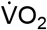 was confirmed visually. Energy expenditure (in mlO_2_/kg/min) of walking was determined by averaging 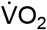 during the last two minutes of each trial. Energy cost (in mlO_2_/kg/m) was calculated by dividing energy expenditure by walking speed. Relative aerobic load was defined as energy expenditure divided by 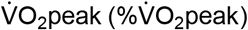 and 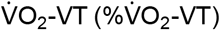 respectively.

### Statistical analysis

Data were analyzed using SPSS (IBM SPSS Statistics version 25.0, IBM Corp.) and checked for normality using visual inspection, skewness and kurtosis inspection and the Kolmogorov-Smirnov and Shapiro-Wilk tests. When the assumption of normality was violated, non-parametric equivalents of statistical tests were used. A p-value of <0.05 was considered significant.

Since previous studies showed that the relationship between speed and energy cost in individuals post-stroke depends on the level of impairment[15], the group was split based on preferred walking speed: a less impaired group, (PWS >0.8 m/s) and a more impaired group (PWS ≤0.8 m/s)[33].

The potential difference between groups in relative aerobic load (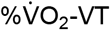 and 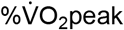) and energy cost at PWS was assessed with two, one-way ANOVA’s. Significant main effects were followed up by post-hoc Bonferroni testing.

To describe the relationships between walking speed and energy cost and between walking speed and relative aerobic load for all groups, Generalized Estimating Equations (GEE) were used. GEE is a group based approach that takes the dependency of observations within individuals over conditions into account. The following equation was used to model the relationship between speed and metabolic parameters:

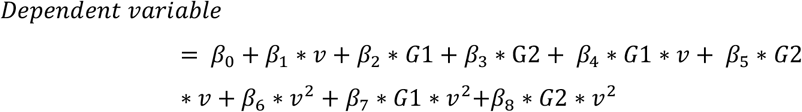

In this equation *v* represents walking speed (m.s^-1^), *G*1=1 represents the more impaired stroke group and *G*2=1 represents the less impaired stroke group. The quadratic component was introduced since previous studies found a partly quadratic relation between speed and both relative aerobic load and energy cost[15,34]. Both main and interaction effects of group and speed were taken into account. Non-significant interaction parameters were removed from the final equations.

## Results

Three out of 45 individuals post-stroke did not finish the protocol: two found the mask too uncomfortable and one found the protocol too tiring. Furthermore, one participant could not perform the protocol within two weeks after the CPET and one participant could not participate due to a fall. Twenty-one post-stroke individuals were allocated to the more impaired group, 19 were allocated to the less impaired group. All 15 able-bodied subjects completed the protocol.

Eleven participants from the more impaired stroke group did not reach 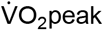 according to our criteria and were therefore not included in the analysis of 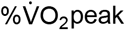. All participants did reach VT and were included in the analysis of 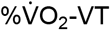. No differences in age, gender or BMI between groups were found (Table 1). As expected, the aerobic capacity of individuals post-stroke was significantly lower than that of able-bodied. No significant difference in aerobic capacity was found between stroke groups.

**Table 1.**
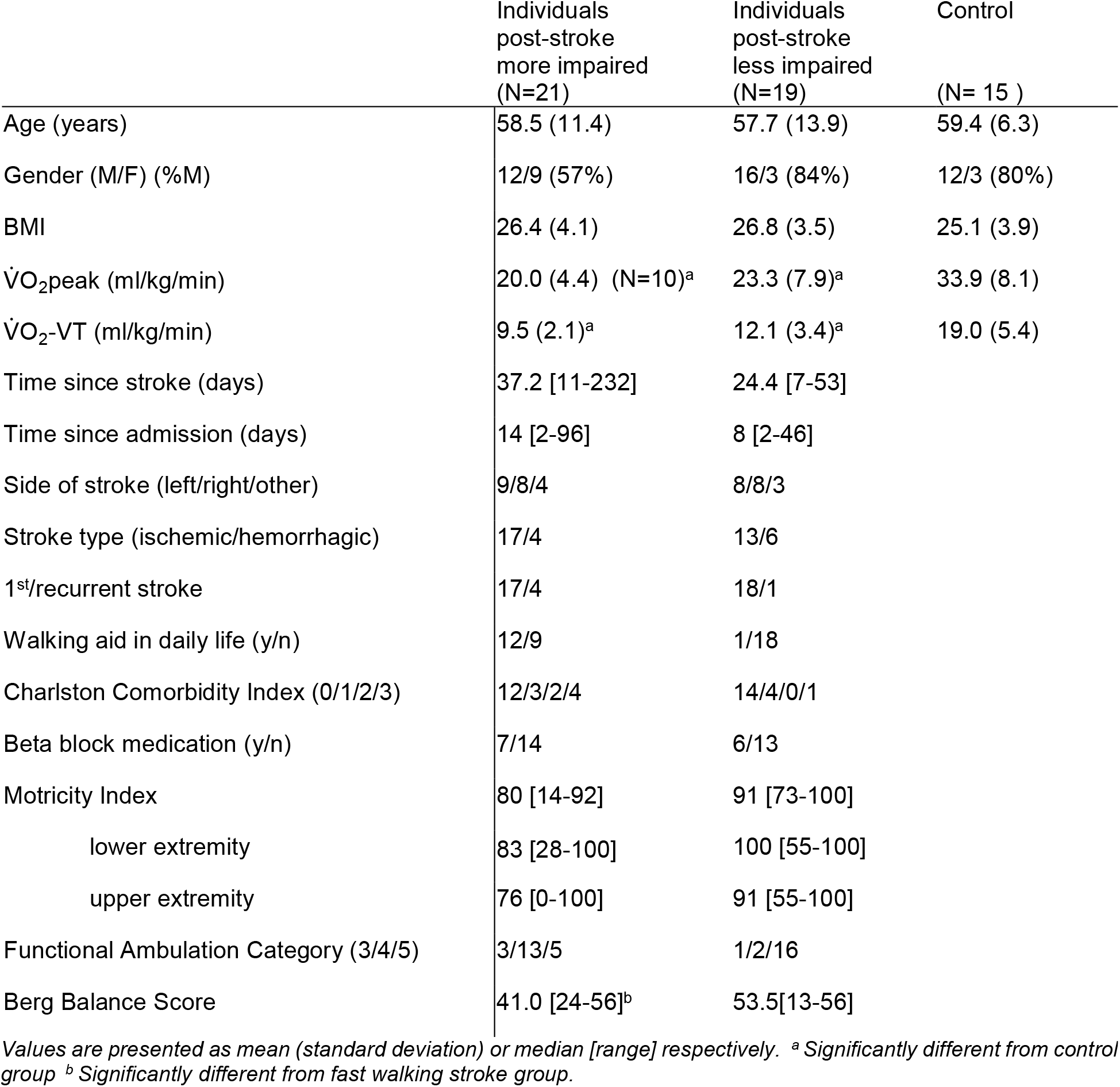
Group characteristics

|  | Individuals<br>post-stroke<br>more impaired<br>(N=21) | Individuals<br>post-stroke<br>less impaired<br>(N=19) | Control<br>(N= 15 ) |
| --- | --- | --- | --- |
| Age (years) | 58.5 (11.4) | 57.7 (13.9) | 59.4 (6.3) |
| Gender (M/F) (%M) | 12/9 (57%) | 16/3 (84%) | 12/3 (80%) |
| BMI | 26.4 (4.1) | 26.8 (3.5) | 25.1 (3.9) |
| $\dot{V}O_2$peak (ml/kg/min) | 20.0 (4.4) (N=10)<sup>a</sup> | 23.3 (7.9)<sup>a</sup> | 33.9 (8.1) |
| $\dot{V}O_2$-VT (ml/kg/min) | 9.5 (2.1)<sup>a</sup> | 12.1 (3.4)<sup>a</sup> | 19.0 (5.4) |
| Time since stroke (days) | 37.2 [11-232] | 24.4 [7-53] |  |
| Time since admission (days) | 14 [2-96] | 8 [2-46] |  |
| Side of stroke (left/right/other) | 9/8/4 | 8/8/3 |  |
| Stroke type (ischemic/hemorrhagic) | 17/4 | 13/6 |  |
| 1<sup>st</sup>/recurrent stroke | 17/4 | 18/1 |  |
| Walking aid in daily life (y/n) | 12/9 | 1/18 |  |
| Charlston Comorbidity Index (0/1/2/3) | 12/3/2/4 | 14/4/0/1 |  |
| Beta block medication (y/n) | 7/14 | 6/13 |  |
| Motricity Index | 80 [14-92] | 91 [73-100] |  |
| lower extremity | 83 [28-100] | 100 [55-100] |  |
| upper extremity | 76 [0-100] | 91 [55-100] |  |
| Functional Ambulation Category (3/4/5) | 3/13/5 | 1/2/16 |  |
| Berg Balance Score | 41.0 [24-56]<sup>b</sup> | 53.5[13-56] |  |

Values are presented as mean (standard deviation) or median [range] respectively. <sup>a</sup> Significantly different from control group <sup>b</sup> Significantly different from fast walking stroke group.

### Relative load and energy cost at PWS

In line with our hypothesis, PWS was significantly different between groups (χ^2^=42.9, p<0.000, Table 2). PWS of the control group was higher than both stroke groups (U=0.0, p<0.000 and U=42.5, p<0.001 respectively). Also in line with our hypothesis, we found a main effect of group on both 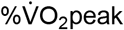 at PWS (F(2,20.8)=8.571, p=0.002) and 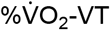 at PWS (F(2,33.9)=14.146, p=0.000). The 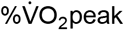 for both the more impaired (50.2[14.4] %) and the less impaired stroke group (51.7[16.8] %) was significantly higher than that of the control group 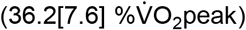, (p=0.007 and p=0.047 respectively). The same was true for 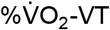; both the more impaired 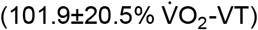 and the less impaired stroke group 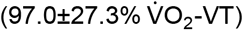 had a significantly higher relative load than the control group 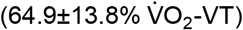, (both p=0.000). No significant differences in relative load at PWS were found between stroke groups.

**Table 2.**
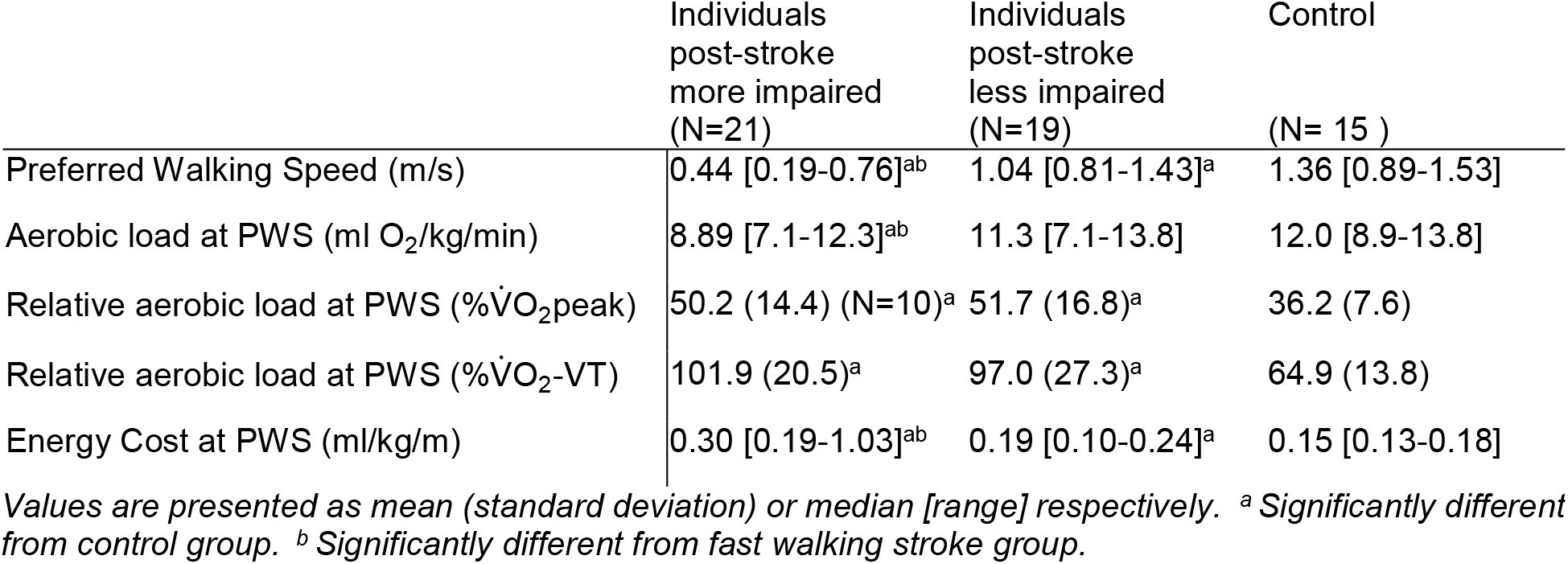
Relative load and energy cost at PWS

As hypothesized, energy cost at PWS was significantly higher in the more impaired stroke group (0.30[.19-1.03] ml/kg/m), compared to the less impaired group (0.19[0.10-0.24] ml/kg/m, p=0.000) and the control group (0.15[0.13-0.18] ml/kg/m, p=0.000, Table 2). Furthermore, energy cost at PWS in the less impaired stroke group was significantly higher than in the control group (p=0.000).

### Relative load as a function of walking speed

Relative aerobic load changed curvilinearly with speed in all groups (Figure 1). For 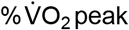, no significant interaction effects of group and speed were found (Table 3). Significant group effects show that 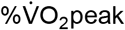 was systematically higher for both stroke groups compared to able-bodied individuals. For 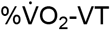, only the interaction between speed and the less impaired group, and speed^2^ and the more impaired group were significant (Table 3).

**Table 3.** GEE results relative load

| Relative Load in $\% \dot{V}O_{2peak}$ | $\beta$ | p-value |
| --- | --- | --- |
| Speed | 21.86 | 0.00* |
| More impaired group | 38.40 | 0.00* |
| Less impaired group | 24.36 | 0.00* |
| Speed<sup>2</sup> | 3.44 | 0.11 |
| Intercept | 2.69 | 0.59 |
| Relative Load in $\% \dot{V}O_{2-VT}$ | | |
| Speed | 9.72 | 0.45 |
| More impaired group | 50.58 | 0.00* |
| Less impaired group | 37.23 | 0.00* |
| Speed<sup>2</sup> | 15.20 | 0.00* |
| More impaired group * Speed | 34.68 | 0.00* |
| Less impaired group * Speed<sup>2</sup> | 7.86 | 0.00* |
| Intercept | 26.58 | 0.01* |

**Figure 1.**
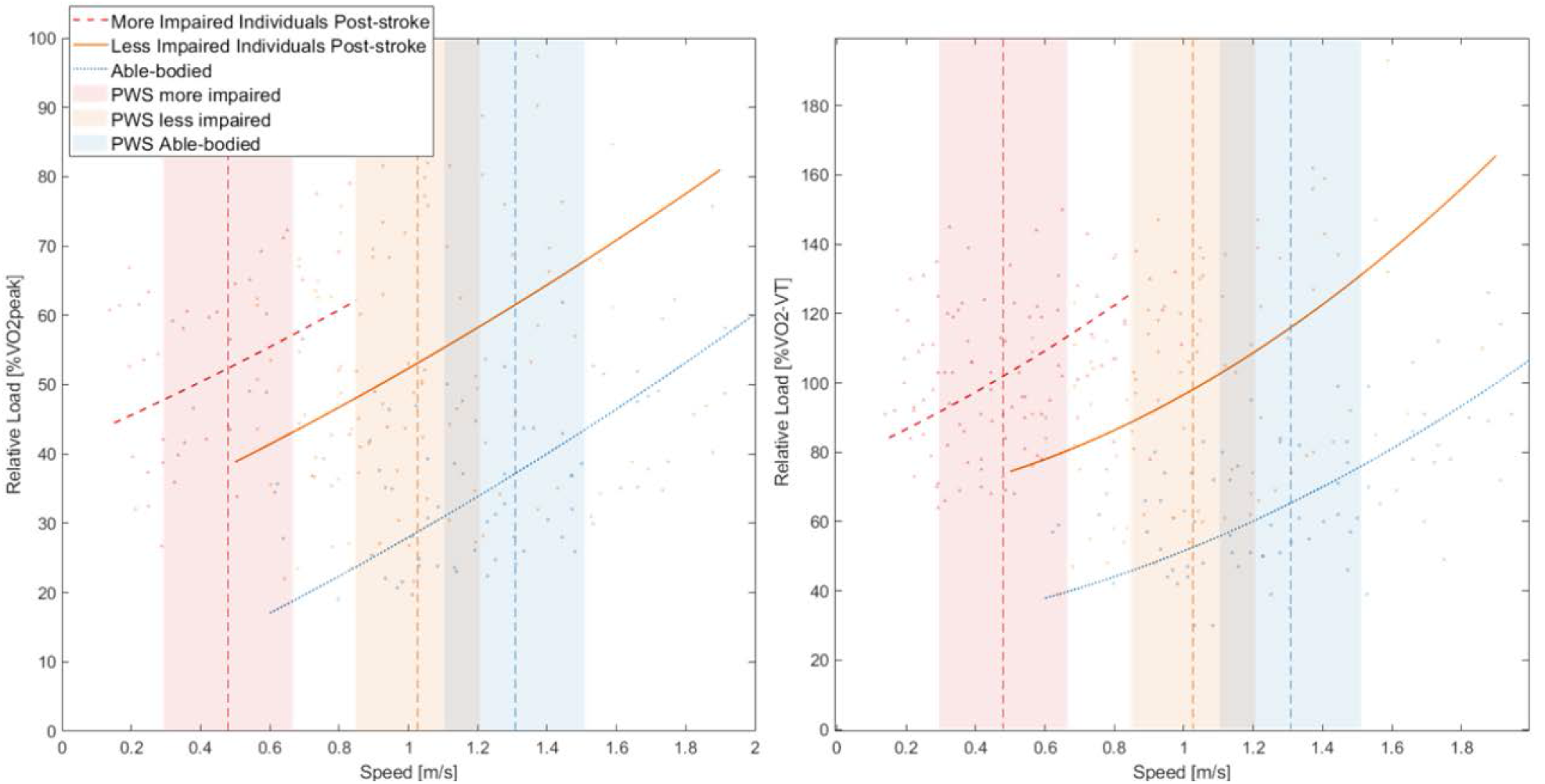
Relative load as a function of walking speed. Relative aerobic load in 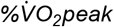 (left panel) and 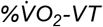 (right panel) in relation to walking speed for all groups. Lines represent the fit according to GEE analysis, which takes the dependency of observations within individuals over conditions into account. For each group, preferred walking speed (PWS) is represented by a dotted vertical line, the shaded area represents the standard deviation of PWS for this group. Individual data points are represented by downward pointing triangles (more-impaired stroke group), upward pointing triangles (less-impaired stroke group) and dots (control subjects) respectively. Note that the range of walking speeds differed between participants and groups since it depended on the PWS of the individual.

### Energy cost as a function of walking speed

At similar speeds, energy cost was higher for both stroke groups compared to able-bodied individuals (Figure 2). Energy cost decreased with speeds above PWS for both stroke groups and this effect was larger for the more impaired group. Able-bodied PWS was close to predicted minimal energy cost.

**Figure 2.**
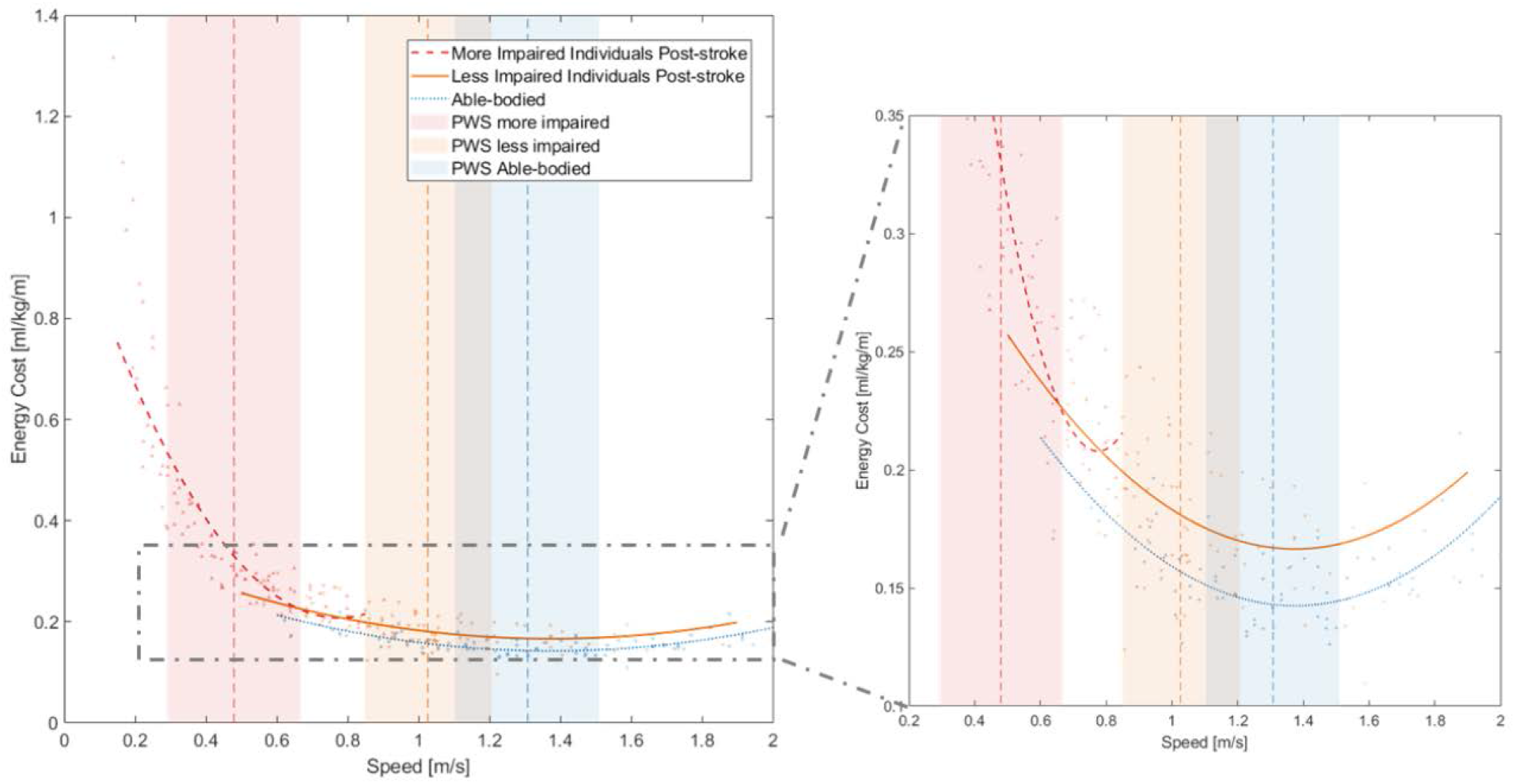
Energy cost as a function of walking speed. Energy Cost in relation to walking speed for all groups. Lines represent the relationship for each group according to GEE analysis, which takes the dependency of observations within individuals over conditions into account. For each group, preferred walking speed (PWS) is represented by a dotted vertical line, the shaded area represents the standard deviation of PWS for this group. Individual data points are represented by downward pointing triangles (more-impaired stroke group), upward pointing triangles (less-impaired stroke group) and dots (control subjects) respectively. Note that the range of walking speeds differed between participants and groups since it depended on the PWS of the individual.

No significant interaction effects with speed and speed^2^ were found for the less impaired group (Table 4).

**Table 4.** GEE results energy cost

| Variable | $\beta$ | p-value |
| --- | --- | --- |
| Speed | -0.326 | 0.000* |
| More impaired group | 0.680 | 0.000* |
| Less impaired group | 0.024 | 0.250 |
| Speed<sup>2</sup> | 0.118 | 0.000* |
| More impaired group * Speed | -1.836 | 0.000* |
| More impaired group * Speed<sup>2</sup> | 1.276 | 0.000* |
| Intercept | 0.366 | 0.000* |

## Discussion

The goal of this study was to identify to what extent relative aerobic load and energy cost could impact walking ability of individuals post-stroke. To this end, we compared relative aerobic load and energy cost of walking between individuals post-stroke and able-bodied individuals.

As hypothesized, we found that relative aerobic load at PWS was higher for individuals post-stroke compared to able-bodied individuals, even though PWS was lowered in individuals post-stroke. At PWS, both more and less impaired individuals post-stroke walked at relative aerobic load values around 100% of ventilatory threshold and 50% of peak aerobic capacity. These values coincide with thresholds for exercise intensity that can be sustained over longer time intervals[4,22]. For able-bodied individuals, relative aerobic load at PWS stayed well below such thresholds. The GEE model reveals that when individuals post-stroke would increase their speed to the PWS of able-bodied, this would lead to unsustainable relative loads of 120%, and 166% of ventilatory threshold for the less and more impaired group respectively (Figure 1). On the other hand, to decrease relative load to values of able-bodied 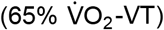, both groups would virtually stand still. Hence, it can be argued that, individuals post-stroke select a PWS that allows them to walk as fast as possible without surpassing acceptable relative load levels.

In line with our hypothesis, energy cost at PWS was higher for individuals post-stroke and highest in the more-impaired stroke group. Likely, this higher energy cost is partly due to the fact that the speed-energy cost curve is elevated for stroke patients, indicating a less efficient walking pattern. This is in accordance with previous studies showing that at equal speed, energy cost is higher post-stroke than in able-bodied individuals[8,16]. However, the high energy cost at PWS appears to be caused mostly by the fact that individuals post-stroke walked below their optimum speed concerning energy cost. Hence, while reducing walking speed limits relative load and prevents fatigue, this comes at the expense of increased energy cost, which aggravates the mismatch between aerobic capacity and aerobic energy demand.

It has often been argued that people select their preferred walking speed such that energy cost is minimized[11,12,14]. It seems that when the cardiovascular system is not a limiting factor, as in able-bodied individuals, walking speed might indeed be optimized for energy cost. However, when the relative load of walking is high, as a consequence of low aerobic capacity or a high energy demand, walking speed seems to be lowered to limit the strain on the cardiorespiratory system at the expense of a higher energy cost. This has previously been observed for individuals with a lower limb amputation[34].

Both increasing aerobic capacity, by exercise training, or decreasing the aerobic load of walking, by improving efficiency, would decrease the relative aerobic load of walking. This would allow faster walking at similar relative load and decreased energy cost. Macko et al.[19], reported the effect of an aerobic treadmill training program of six months for individuals post-stroke in the chronic phase. After training, the relative aerobic load of walking decreased from 62% to 50% 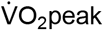, while walking speed increased. This indicates that patients did not only reduce relative aerobic load, but also exploited the training effect to increase movement speed. Likely, they walked with a lower energy cost at the increased walking speed. However, it is unclear from these results to what extent an improved economy, or an improved aerobic capacity contributed to reduced relative load and increased speed. Nevertheless, it seems likely that improving aerobic capacity would make it possible to sustain a higher walking speed, which would in turn lead to a lower energy cost. Thus, for patients who walk at a high relative aerobic load and concomitant high energy cost, it might be worthwhile to consider training aerobic capacity when addressing walking ability.

### Study limitations

Since this study aimed to compare physiological parameters at and around PWS, the range of speeds at which subjects walked differed substantially between groups. Therefore, we could not directly compare energy cost and relative aerobic load at the low, and high end of the speed range between groups. Furthermore, the imposed second order model on the relation between energy cost and walking speed did not fit this relationship for the more-impaired group very well. Therefore, the regression model for this group, especially the predicted minimum should be interpreted with caution. Advisably, future studies should impose equal speed ranges over all groups to improve comparisons.

A strength of this study is that we measured aerobic capacity via the gold standard: a maximal cardiopulmonary exercise test with standardized criteria for maximal effort. This led to a more reliable estimate of relative aerobic load compared to studies using prediction equations, and to a more complete estimate of the strain of walking compared to studies that only report absolute aerobic load. Furthermore, we included VT in our analyses of relative load. VT was attained by all subjects, also those who did not reach maximal aerobic capacity. Moreover, VT might be a more relevant reference for aerobic strain as it represents the exercise intensity that can be sustained for a long duration.

## Conclusion

Individuals post-stroke seem to reduce their walking speed to prevent unsustainable relative aerobic loads. This comes at the expense of reduced economy (i.e. higher energy cost). A higher aerobic capacity would decrease the relative aerobic load of walking, which would make it possible to increase walking speed and thereby improve economy. Therefore, when aiming to improve walking ability in individuals post-stroke, it is important to consider training aerobic capacity.

## Data Availability

N/A

## Acknowledgements

The authors would like to thank Ilona Visser, Bastiaan Vader, Richard Fickert, Jan-Willem Dijkstra, Tijs van Bezeij, Maarten Tolsma en Feikje Riedstra for their assistance in data collection.

